# The risk factors of COVID-19 in a longitudinal population-based study

**DOI:** 10.1101/2021.04.12.21255369

**Authors:** Hozhabr Jamali Atergeleh, Mohammad Hassan Emamian, Shahrbanoo Goli, Marzieh Rohani-Rasaf, Hassan Hashemi, Akbar Fotouhi

**Affiliations:** Student Research Committee, School of Public Health, Shahroud University of Medical Sciences, Shahroud, Iran; Ophthalmic Epidemiology Research Center, Shahroud University of Medical Sciences, Shahroud, Iran; Department of Epidemiology, School of Public Health, Shahroud University of Medical Sciences, Shahroud, Iran; Noor Ophthalmology Research Center, Noor Eye Hospital, Tehran, Iran; Department of Epidemiology and Biostatistics, School of Public Health, Tehran University of Medical Sciences, Tehran, Iran

## Abstract

The present longitudinal study aims to investigate the risk factors for getting COVID-19 in a population aged 50 to 74 years. Data were collected from Shahroud Eye Cohort Study and the electronic system of COVID-19 in Shahroud, northeast Iran. Participants were followed for about 13 months and predisposing factors for COVID-19 infection were investigated using log binomial model and by calculation of relative risks. From the beginning of the COVID-19 outbreak in Shahroud (February 20, 2020) to March 26, 2021, out of 4394 participants in the Eye Cohort Study, 271 (6.1%) were diagnosed with COVID-19 with a positive Reverse Transcription Polymerase Chain Reaction test on two nasopharyngeal and oropharyngeal swabs. Risk factors for getting COVID-19 were included male gender (Relative Risk (RR) = 1.51; 95% Confidence Intervals (CI), 1.15-1.99), BMI over 25 (RR = 1.03; 95% CI, 1.01-1.05) and diabetes (RR = 1.31; 95% CI, 1.02-1.67). Also, smoking (RR = 0.51; 95% CI, 0.28-0.93) and education (RR = 0.95; 95% CI, 0.92-0.98) had reverse associations. In conclusion men and diabetic patients and those who have BMI over 25, should be more alert to follow the health protocols related to COVID-19 and priority should be given to them considering COVID-19 vaccination.

## Introduction

COVID-19, which was first reported from China in 2019 and became a pandemic within a few months, is a threat to human society and has challenged all aspects of human life. As of 5 April 2021, more than 130 million people have been infected with the disease worldwide, and caused more than 2800000 deaths. In Iran, more than 1900000 cases have been identified and more than 62000 deaths have been reported [1]. One of the characteristics of this disease is its rapid transmission, which spread it to all parts of the world in a short time; therefore, it is necessary to identify ways to prevent and treat this disease to restore normal conditions to human societies. One of the main ways to deal with COVID-19 infection is to identify the its predisposing risk.

Various studies have investigated the risk factors for COVID-19 infection, which identified risk factors like age, sex, race, occupation, level of education and social and economic status [2-6] and underlying diseases such as cardiovascular disease, Chronic kidney disease and hypertension [7]. Most of this information is obtained from cross-sectional studies that may have biases related to this type of study. Longitudinal studies, due to their design, can provide more accurate information. In the present study as a longitudinal one, these risk factors were investigated. Other longitudinal studies have been investigated COVID-19 in Iran [8, 9], but with lower follow up periods, and with focus on risk factors of severe disease and death. This study aims to investigate risk factors for getting COVID-19 infection after 13 months follow up of people.

## Methods

Shahroud Eye Cohort Study began with the invitation of 6311 people in 2009, of which 5190 people entered the study [10]. The study was conducted in three phases at five-year intervals in adult population of Shahroud, northeast Iran. Many demographic variables such as gender, age, marital status, education, occupation and also smoking, underlying diseases, blood group, body mass index, blood pressure and clinical tests were recorded in Eye Cohort Study. The third phase of this study began in 2019 and ended before the beginning of the COVID-19 outbreak in Iran with 4394 participants aged 50 to 74. In this study, participants in the third phase of Eye Cohort Study were followed for COVID-19 infection over a 13 months period.

The required data were extracted from the Eye Cohort Study Database and the electronic system for recording and following up cases of COVID-19. The data from the two sources were combined to identify the participants in the Shahroud eye cohort study who had COVID-19 symptoms and underwent the COVID-19 diagnostic tests, and finally these data were organized.

The definitive cases were those who referred to health centers with symptoms of the disease and whose Reverse Transcription Polymerase Chain Reaction (RT-PCR) test was reported to be positive for nasopharyngeal and oropharyngeal swabs. Widowers and divorcees were considered as single. Those whose systolic blood pressure was ≥140 mm Hg and / or their diastolic blood pressure was ≥ 90 mm Hg, or those who were taking antihypertensive medications were considered as hypertensive. People with diabetes were those with Fasting Plasma Glucose ≥ 7 mmol / L and / or people on Glucose-lowering medicines. People with Dyslipidemia were those with Triglyceride ≥ 2.26 mmol / L, and / or Cholesterol ≥ 6.21 mmol / L, and / or HDL-C < 1.03 mmol / L, and / or using lipid-lowering medications.

For statistical analysis, mean and standard deviation or relative frequencies of variables were calculated in both cases and non-cases who were categorized based on RT-PCR test results. The association between outcome of interest (COVID-19 patients) and studied variables were examined using independent samples t-test or chi-square test. The simple and multiple binomial regression models were used to calculate unadjusted and adjusted relative risk for getting COVID-19.

The Shahroud Eye Cohort Study and this study were approved by the ethics committee of Shahroud University of Medical Sciences. All participants signed written informed consents.

## Results

From the beginning of the COVID-19 outbreak in Shahroud (February 20, 2020) to March 26, 20201, 271 people (6.1%) were diagnosed with COVID-19 infection with RT-PCR test among the 4394 participants of Shahroud eye cohort study. Table 1 shows the distribution of participants’ demographic information and their risk factors exposure, categorized by the results of RT-PCR test.

**Table 1:**
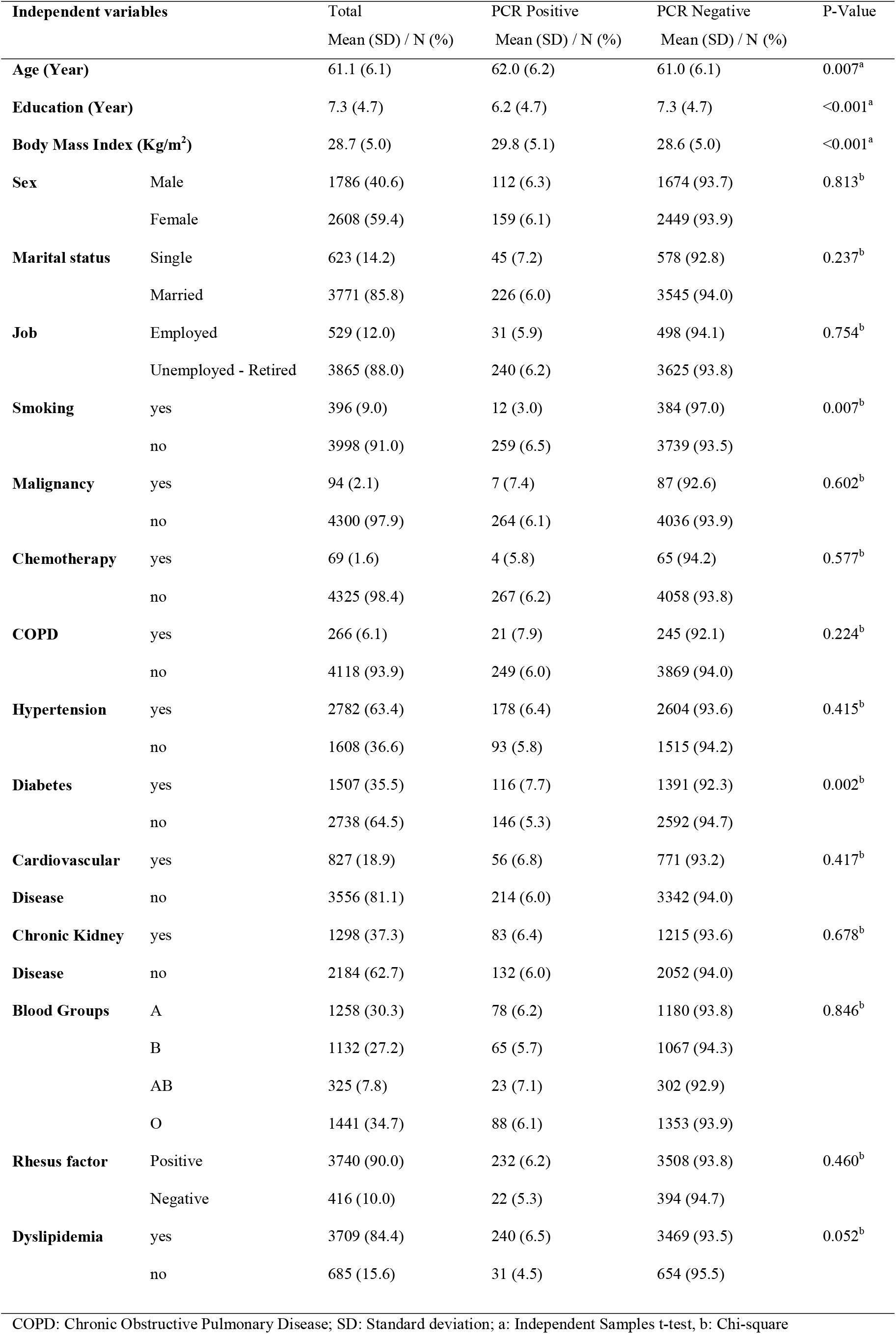
Demographic characteristics and past medical history of participants by Polymerase Chain Reaction (PCR) test results, Shahroud, Iran.

**Table 2:** The association of demographic variables and past medical conditions with COVID-19 in Log Binominal Regression models, Shahroud, Iran.

| Independent Variables | Simple |  |  | Multiple |  |  |
| --- | --- | --- | --- | --- | --- | --- |
|  | RR | 95% CI | P-Value | RR | 95% CI | P-Value |
| Age | 1.02 | 1.00-1.04 | 0.007 | 1.01 | 0.99-1.03 | 0.316 |
| Male Sex | 1.02 | 0.81-1.30 | 0.813 | 1.51 | 1.15-1.99 | 0.003 |
| Being married | 0.82 | 0.60-1.12 | 0.236 |  |  |  |
| Job | 0.94 | 0.65-1.35 | 0.754 |  |  |  |
| Having Malignancy | 1.21 | 0.58 -2.49 | 0.600 |  |  |  |
| COPD<sup>2</sup> | 1.30 | 0.85-2.00 | 0.222 |  |  |  |
| Smoking | 0.46 | 0.26-0.82 | 0.009 | 0.51 | 0.28-0.93 | 0.030 |
| Under Chemotherapy | 0.93 | 0.36-2.44 | 0.898 |  |  |  |
| Hypertension | 1.10 | 0.86-1.41 | 0.416 |  |  |  |
| Diabetes | 1.44 | 1.14-1.82 | 0.002 | 1.31 | 1.02-1.67 | 0.031 |
| CVD<sup>3</sup> | 1.12 | 0.84-1.49 | 0.416 |  |  |  |
| CKD<sup>4</sup> | 1.05 | 0.81-1.38 | 0.678 |  |  |  |
| Dyslipidemia | 1.42 | 0.99-2.05 | 0.055 |  |  |  |
| Education (year) | 0.95 | 0.92-0.97 | <0.001 | 0.95 | 0.92-0.98 | 0.002 |
| BMI<sup>5</sup> | 1.04 | 1.02-1.06 | <0.001 | 1.03 | 1.01-1.05 | 0.001 |
| Positive Rh<sup>6</sup> | 0.85 | 0.55-1.30 | 0.462 |  |  |  |
| Blood Groups |  |  |  |  |  |  |
| AB (reference) | 1 |  |  |  |  |  |
| A | 0.87 | 0.55-1.37 | 0.564 |  |  |  |
| B | 0.81 | 0.51-1.28 | 0.372 |  |  |  |
| O | 0.86 | 0.55-1.34 | 0.514 |  |  |  |

CI: Confidence Interval; COPD: Chronic Obstructive Pulmonary Disease; CVD: Cardiovascular
Disease; CVD: Chronic Kidney Disease; BMI: Body Mass Index (kg/m<sup>2</sup>); RH: Rhesus (Rh) factor

The mean age of the participants was 61.1 years and most of the participants (59.4%) were female. Dyslipidemia (84.4%) and hypertension (63.4%) were the most common among the underlying diseases. The highest frequency of blood groups was in group O (34.7%) and the lowest frequency was in group AB (7.8%).

The mean age of PCR-positive cases (62.0 year) was higher than that of negative cases (61.0 year). Also, the prevalence of smoking was higher among PCR-negative individuals. Mean BMI as well as the frequency of dyslipidemia were higher in PCR positive cases. The mean years of education in PCR-negative cases were higher than PCR-positive cases.

In simple binomial regression models, the relation between each independent variable and the COVID-19 was calculated. Among the studied variables, age, diabetes, smoking, BMI and education had a significant relation with the outcome.

In multiple binomial regression model, all variables that were significant in simple models were entered into the model. In this model, the relative risk of getting COVID-19 for males was 1.51 (95% CI, 1.15-1.99). Higher BMI also increased the risk of COVID-19 (RR = 1.03; 95% CI, 1.01-1.05) and diabetes was associated with an increased risk of the disease (RR = 1.31; 95% CI, 1.02-1.67). Smoking and higher level of education reduced the risk of COVID-19 (RR = 0.51; 95% CI, 0.28-0.93 and RR = 0.95; 95% CI, 0.92-0.98, respectively).

## Discussion

Identifying risk factors and high-risk groups for COVID-19 are of particular importance for controlling and reducing the burden of disease in communities. In the present study male gender, high BMI, smoking, low education, and diabetes were associated with higher risk of COVID-19 infection.

Male gender has been identified as a risk factor for severe morbidity and death from COVID-19 in other studies [11, 12]. In a meta-analysis of 57 studies, the prevalence of COVID-19 was higher in men [13]. The reason for this intergender difference can be explained in the molecular and hormonal differences between the two gender as well as the behaviors of the males. The higher prevalence of employment among men (83.2% in the present study) as well as more risky behaviors and non-compliance with healthy and nutritional recommendations can be reasons for this increased risk. Drinking alcohol and smoking are also other factors in making men more susceptible to the diseases, although in the current study smoking was a protective factor.

There are other studies that have shown smoking a protective factor and showed that smokers have a lower risk of developing COVID-19 infection [14, 15] and even worse outcomes of the disease [16-19], but other studies have indicated worsened outcomes and higher death rate from COVID-19 among smokers [20-24]. Some consider the role of tobacco mosaic virus (TMV) in antibody production and interferon secretion as a mechanism that tobacco use protect people against COVID-19 infection [17]. Even current smokers were more protected against COVID-19 than former smokers [25]. There is a huge warning agenda against smoking risk and its relation to COPD in media, so perhaps smokers have more healthy behaviors, such as social distance and wearing a mask, because they feel more risk for getting COVID-19. There are also evidences that show the desire to quit smoking and not smoking in closed environments has increased after the COVID-19 pandemic [26]. Most of these studies are cross-sectional, and more powerful studies should be conducted longitudinally to examine the true effect of smoking on COVID-19.

A study in India showed that the higher BMI is a risk factor for COVID-19 infection [27], the same in our study, each unit increase in BMI increased the risk of COVID-19 by three percent. High body mass index is one of the risk factors for many diseases and being overweight and obese disrupts the immune system [28]. Obese or overweight people may even have more occupationally high-risk behaviors and follow less prevention protocols for COVID-19. Even being overweight can affect the outcome of an infection and studies show that a higher BMI can be a risk factor for the infection, worsening the condition of a person with COVID-19, and a predictor of ICU admission and even death [29-31].

Diabetes was another risk factor for getting COVID-19 in current study. Other study showed no difference in COVID-19 infection prevalence between diabetic population and general population [32], while diabetes was a predictor of ICU admission and death in infected patients [33]. Weak immune system in diabetic patients put them at greater risk for various infections [34, 35], and the same can be true in case of COVID-19. COVID-19 infection can even cause ketoacidosis in diabetic patients [36] and due to electrolyte loss, it is more difficult to control infection in them and this increases the risk of death.

Another factor that was examined in the present study and found to be a protective factor was the level of education. We found that each year increase in the level of education reduces the risk of COVID-19 by five percent. The results of some studies show that a higher level of knowledge about COVID-19 disease, which has been associated with the level of education of individuals, is a strong predictor of the disease [2, 37]. It seems that various factors can affect this protective effect. Better economic status of people with higher education, more awareness of these people due to more reading and more access to the media and adhering social distance and health protocols, can be reasons for the protection role of education against COVID-19 infection.

High blood pressure and being employed are among studied variables that were not associated with COVID-19 in current study but have been identified as a risk factor for COVID-19 in other studies [2, 7]. The reason may be high prevalence of hypertension [38] and unemployment in this study. Having malignancy has not been identified as a risk factor for COVID-19 in current study which may be the result of small number of participants with malignancy and low statistical power. Studies show that people have a different chance of developing COVID-19 based on their blood groups and RH types [39-41], but this was not observed in our study. The differences in study design, age distribution of participants and epidemic severity may be the reasons for different results regarding blood groups. Finally older age, which has been identified as a risk factor for COVID-19 [4], was not significant in current study, which may be due to the limited age range and high mean age in this study.

Longitudinal design and high sample size were the strengths of this study. However, this study only examined people aged 50 to 74 years and its results may not be generalized to other age groups. In some subgroups (Participants with malignancy or under chemotherapy) we also had low sample size that may affect the results. Another limitation of this study is the lack of information about the level of knowledge and adherence to health protocols among the participants.

## Conclusion

The results of this longitudinal study showed that being male, having a higher BMI and being diabetic can increase the risk of COVID-19 infection among the population of 50 to 74 years. Also, in this age group having higher education had a protective role against this disease. The lower risk of COVID-19 among smokers needs to be examined more closely to determine whether this is related to smoking or their behavioral patterns. High risk groups should be informed more about COVID-19 and also prioritize them in vaccination programs.

## Data Availability

Data will be available on request

## Acknowledgment

The present study is a part of thesis for master degree (No: 200084). Shahroud Eye Cohort Study was supported by Shahroud University of Medical Sciences (Grant Number: 9826) and Noor Eye Hospital.

